# The 2024 International Consensus Reference Standard for Urinary Tract Infection (UTI) Research fails in Neurogenic Bladder without UTI symptoms

**DOI:** 10.64898/2026.07.10.26357767

**Authors:** Rochelle E. Tractenberg, Suzanne L. Groah

## Abstract

**Background:** The 2024 international reference standard for urinary tract infection (UTI) research scores four domains - symptoms and signs, systemic criteria, pyuria, and culture - to classify samples as No UTI, Possible UTI, Probable UTI, or Definite UTI. It identifies spinal cord injury (SCI) as a condition of impaired symptom perception, and states that catheter-associated UTI requires a separate standard. We applied it to confirmed-asymptomatic samples from adults with neurogenic lower urinary tract dysfunction (NLUTD) due to spinal cord injury or disease (SCI/D) who use intermittent catheterization (IC).

**Methods:** The reference standard was applied to 222 samples from 96 adults with NLUTD due to SCI/D using IC, for all of which the participant confirmed, using the Urinary Symptom Questionnaire for Neurogenic Bladder–Intermittent Catheter (USQNB-IC), that they had been asymptomatic at sampling and for 72 hours prior. Because no participant was febrile and no blood markers are drawn in this population, the systemic-criteria domain scored zero for every sample; the reported classifications are therefore a floor. Pyuria was scored under conservative and inclusive interpretations of categorical urinary white blood cell (WBC) bins.

**Results:** Under the conservative interpretation, 38.7% of samples were classified No UTI, 22.5% Possible UTI, and 38.7% Probable UTI; under the inclusive interpretation, 8.1% No UTI, 40.5% Possible UTI, and 51.4% Probable UTI. No sample reached Definite UTI, a structural consequence of the empty systemic domain. Of the samples that met the pyuria entry threshold, 136 of 136 (conservative) and 204 of 206 (inclusive) were classified Possible or Probable UTI: meeting the entry threshold determined the classification.

**Conclusions:** The consensus reference standard classifies 61–92% of confirmed-asymptomatic NLUTD-IC samples as Possible or Probable UTI, and 39–51% as Probable UTI - a floor estimate that could only rise if blood markers were available. This confirms the framework’s own prediction that a separate standard is needed for populations with altered symptom expression and baseline-positive urinary markers.

**Research in context:** *Evidence before this study:* Urinary tract infection (UTI) is the most common secondary condition and infectious cause of hospitalisation among people with neurogenic lower urinary tract dysfunction due to spinal cord injury or disease, and it is over-diagnosed and over-treated in this population. Diagnosis has long rested on laboratory thresholds - pyuria and bacteriuria - despite repeated demonstrations that both are common in people who do not have or develop symptoms, and that neither is associated with symptom status in those who catheterize. In 2024 an international multidisciplinary Delphi consensus published a reference standard for UTI research that scores four domains, states that it systematically addressed all issues of diagnosis and nomenclature for research purposes, and extends explicitly to populations unable to perceive or express lower urinary tract symptoms, naming spinal cord injury as one such population. That standard has not, to our knowledge, been applied to people in this population whose symptom status was known, and no published study has examined whether the reference standard can be applied based on the results in routine clinical laboratory reports.

*Added value of this study:* We applied the reference standard to 222 urine samples from 96 community-dwelling adults with neurogenic lower urinary tract dysfunction who use intermittent catheterisation, each of whom confirmed, using an instrument validated in this population, that they had been asymptomatic for the 72 hours before the sample was taken. The standard assigns one point toward a UTI classification to individuals with spinal cord injury without requiring them to endorse any specific symptom, and in this group of asymptomatic individuals, between 61% and 92% of their samples were classified as Possible or Probable UTI. The range of misclassification depends on how the pyuria criteria are applied: a large clinical laboratory provides leukocyte counts in bins that overlap the reference standard criteria at the low (0-5 per hpf) and high (40-60 per hpf) ends, whereas a research laboratory might instead provide exactly the values requested.

*Implications of all the available evidence:* A reference standard intended to define UTI for research is intended for selecting participants, adjudicating patient and research outcomes, and auditing practice. If it classifies the majority of samples from people who report no symptoms as Possible or Probable UTI, it will inflate apparent infection rates, systematically misclassify controls, and give the appearance of evidence to treatment decisions that the underlying data do not support. This is demonstrated within a population already subject to substantial antimicrobial over-use, those with neurogenic lower urinary tract dysfunction due to spinal cord injury or disease. Two remedies follow directly. The capability question should be answered from the patient, using instruments that are validated for the patient’s primary method of bladder management, rather than from the diagnosis; and any standard intended for routine as well as research settings should be scoreable from the results laboratories actually issue, with explicit instructions for categorical bins that straddle its thresholds and for results that are categorical rather than quantitative.

## Introduction

Urinary tract infection (UTI) is the most common outpatient infection worldwide and the leading infectious cause of hospitalisation among people with spinal cord injury or disease (SCI/D) and neurogenic lower urinary tract dysfunction (NLUTD).1–3 These individuals frequently have bacteriuria and pyuria without symptoms, so the markers used to diagnose UTI in neurologically intact people - culture, urinalysis and dipstick - reflecting a different baseline rather than infection.4–6 The consequence is systematic misdiagnosis: asymptomatic bacteriuria is treated as UTI, contributing to inappropriate prescribing, resistance and unnecessary hospitalization.7 That these markers are unreliable for UTI diagnosis in this population is already established;4,5,8 the gap this paper addresses is not one of missing knowledge but of its implementation.

In 2024, Bilsen and colleagues published an international, multidisciplinary Delphi consensus reference standard for UTI research.9 The framework scores four domains - symptoms and signs, systemic criteria, pyuria, and culture - in order to classify each sample as representing No UTI (0–2 points), Possible UTI (3–4), Probable UTI (5–7), or Definite UTI (≥8). The standard achieved 94% consensus among 46 international experts.

The framework identifies SCI as a condition associated with “inability to perceive or express symptoms”, for which it awards one point in the symptoms and signs domain automatically. However, in their discussion, the authors state that catheter-associated UTI “requires a separate reference standard” because symptoms and laboratory interpretation are “even more challenging” there. Both features - altered symptom expression and baseline-positive markers - are well known to characterize individuals with NLUTD due to SCI/D. Despite these caveats the framework is described as having “systematically addressed all issues related to UTI diagnosis and nomenclature”. To our knowledge it has not been empirically tested.

We applied the framework to repeat urine samples from adults with NLUTD due to SCI/D using IC who confirmed they were asymptomatic for 72 hours prior to the sample to determine whether the framework can perform a core task in UTI diagnosis: distinguishing asymptomatic bacteriuria from symptomatic UTI.

## Methods

### Participants and samples

Participants and urinary marker data are described in full elsewhere.6 IRB approval (#1124) and informed consent were obtained. Briefly, community-dwelling adults with NLUTD due to SCI/D who manage their bladders with intermittent catheterisation provided urine samples at scheduled visits. For every sample the participant confirmed, using the Urinary Symptoms Questionaire for Neurogenic Bladder-Intermittent Catheter version (USQNB-IC) - a validated patient-reported outcome instrument for urinary symptoms in this population10 - that they had been asymptomatic at collection and for the 72 hours prior. Participants also confirmed they had abstained from sexual intercourse for 72 hours prior to the sample.

Of 235 samples, 3 were excluded for documented systemic antibiotic exposure at collection, 3 because symptom status could not be adjudicated, and 1 for absent demographic data. A further 6 lacked a urinalysis, which is essential because pyuria is the entry criterion for the framework. This yielded 222 samples from 96 asymptomatic adults (two contributed one sample, 70 two, 16 three, eight four). The USQNB-IC was completed immediately after the sample (time 0) and at 24, 48 and 72 hours afterwards. 192 samples remained asymptomatic at all four assessments and in 30 cases, the individual endorsed at least one symptom within 72 hours. In one case the individual developed a diagnosed UTI within 24 hours of the sample.

### Applying the standard

The framework is a routed algorithm, not just a simple sum of scores. There are two questions preceding scoring where, if the sample ‘exits’ the algorithm, it is classified No UTI. The first question asks whether the patient can perceive or express lower urinary tract symptoms, and names spinal cord injury as an example of inability. A “yes” leads to the symptom question, where a patient without new-onset symptoms is classified No UTI immediately; a “no” bypasses that question entirely and instead awards one point in the symptoms domain for the same inability. Every sample scored one symptom point based on the SCI example at the entry point of the algorithm. Since no individual had blood drawn and all were asymptomatic at the time of sample, every sample scored zero systemic points. This caps the total possible points at 7, making Definite UTI (≥8) unattainable for these samples.

### Pyuria criteria and the two interpretations

The entry threshold is ≥10 leukocytes per µL or ≥5 per hpf; the standard awards 2 points to 10-200 per µL or 5-50 per hpf and 3 points for higher values, subtracting one point from the 2-point band only for women aged ≥65 years. Our laboratory reports leukocytes in ordered categorical bins per hpf, two of which straddle a cut point and two of which are not counts at all. We therefore scored every sample twice: *conservatively*, a bin met a threshold only if its entire range cleared it, so “00-05” was routed to No UTI and “40-60” scored 2; *inclusively*, a bin met a threshold if any part of its range reached it, so “00-05” entered scoring and “40-60” scored 3. The non-count results our laboratory provides are “none seen” and “packed”. These are treated ordinally as the lowest and highest values possible, respectively (according to the laboratory data dictionary). Because the entry threshold is a routing step, the pyuria result determines how many samples the algorithm evaluates at all: 136 of 222 were scored conservatively and 206/222 were scored inclusively. S1 Table presents all of these results.

### Culture scoring

The standard awards 0 points for no growth or culture not performed, 1 for ≥3 organisms, mixed flora or an “other pathogen”, and 3 for a typical uropathogen at ≥10³ CFU/mL for a single catheterized specimen; women aged ≥65 years with the latter result receive 2 rather than 3 points. Culture results reported as mixed flora with no organism named were scored 1, per the standard’s mixed-flora cell (n = 4); cultures performed that grew nothing (no result but a comment, “no growth”) were treated as below the threshold (n = 26); and results reported as “fewer than 10,000 CFU/mL” of a single unspeciated organism were left unscored (n = 3), as were 11 cultures that were never performed. S2 Table summarizes all of these results.

### Classification and analysis

Domain scores were summed and classified as No UTI (0–2), Possible UTI (3–4), Probable UTI (5–7) or Definite UTI (≥8). Beyond classification we audited the instrument, asking of each domain whether it varies, what share of variance it contributes, whether its categorisation is faithful to the underlying measurement, whether components are redundant, whether the scoring resolution is supported by the precision of the measurement, what range is attainable against the nominal range, and whether routing and scoring are separable. Scoring was performed in SPSS v.29 (IBM 2024). Claude (Opus 5, Anthropic) was used to confirm scoring and counts, test and compare different scoring and algorithm applications, generate figures and tables, and to ensure complete and correct reporting in this manuscript.

## Results

Table 1 presents category assignments for all 222 analytic samples, and Figure 1 shows the routing of those samples under both possible answers to the first question.

**Figure 1.**
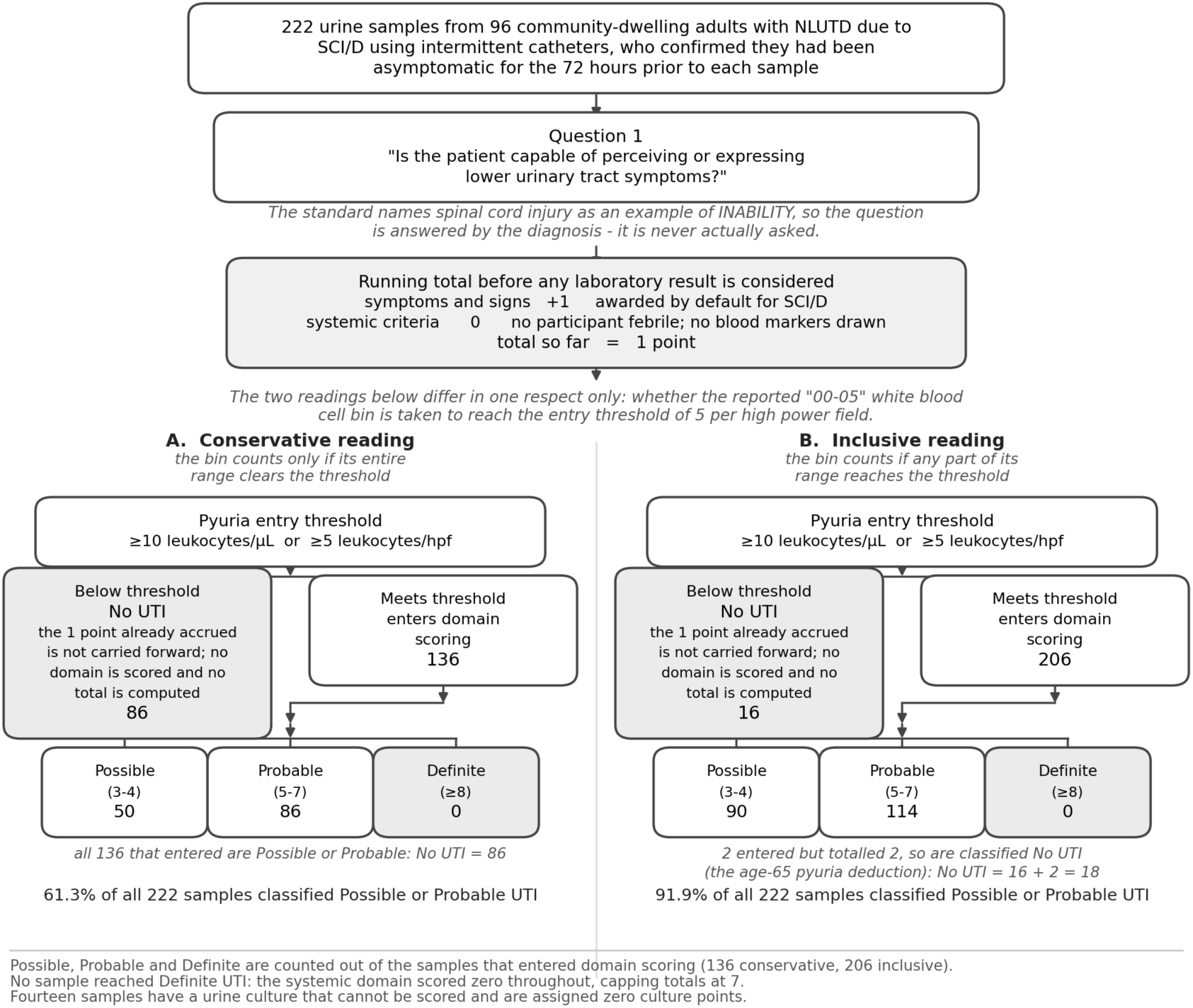
Route taken by 222 confirmed-asymptomatic samples through the reference standard, as the standard directs it be applied to this population. Counts are given as conservative | inclusive interpretation of the white blood cell bin that straddles the pyuria entry threshold. The first question is answered by the diagnosis rather than by assessment, the standard naming spinal cord injury as an example of inability to perceive or express lower urinary tract symptoms; that single answer both removes the symptom-based exit and awards a symptom point, so routing and scoring are not separable. Boxes in the right-hand panel are not steps in the algorithm: they set out what the algorithm does not accommodate, including the validated patient-reported instruments that exist for each method of bladder management. Samples falling below the pyuria entry threshold are classified No UTI without any domain being scored and without a numeric total; this is not a total of zero, and the symptom point those samples carry is never added up. Had the first question been answered from the symptom data available for this cohort, all 222 samples would have been classified No UTI without any domain being scored.

**Table 1.** Bilsen et al. (2024) UTI category assignments for 222 samples from asymptomatic individuals with SCI/D.

| | No UTI<br>(0–2) | Possible UTI<br>(3–4) | Probable UTI<br>(5–7) | Definite UTI<br>($\geq 8$)* |
| --- | --- | --- | --- | --- |
| **Conservative**<br>**(N=222)** | 86 (38.7%) | 50 (22.5%) | 86 (38.7%) | 0 (0%) |
| Asymptomatic<br>72h<br>(n=192) | 76 (39.6%) | 42 (21.9%) | 74 (38.5%) |  |
| Developed ≥1<br>symptom<br>within 72h<br>(n=30) | 10 (33.3%) | 8 (26.7%) | 12 (40.0%) |  |
| **Inclusive**<br>**(N=222)** | 18 (8.1%) | 90 (40.5%) | 114 (51.4%) | 0 (0%) |
| Asymptomatic<br>72h<br>(n=192) | 15 (7.8%) | 79 (41.1%) | 98 (51.0%) |  |
| Developed ≥1<br>symptom<br>within 72h<br>(n=30) | 3 (10.0%) | 11 (36.7%) | 16 (53.3%) |  |

Note. Conservative = a bin was scored above a threshold only if its entire range cleared it ("00-05" failed to clear the ≥5 per hpf threshold and generated "No UTI"; "40-60" failed to clear the ≥50 per hpf threshold and was scored 2 points, or 1 where the age-65 deduction applied). Inclusive = a bin was scored above a threshold if any part of its range reached it ("00-05" cleared the ≥5 per hpf threshold and entered scoring at 2 points, or 1 with that deduction; "40-60" cleared the ≥50 per hpf threshold and was scored 3 points, or 2 with that deduction). All other bins scored identically under both interpretations, "none seen" failed the ≥5 per hpf threshold; "packed" cleared the ≥50 per hpf threshold. Samples not meeting the pyuria entry threshold are classified No UTI without domain scoring and have no numeric total: 86 of the 86 No UTI samples under the conservative interpretation, and 16 of the 18 under the inclusive. The two remaining inclusive samples entered scoring but totalled 2, below the Possible UTI threshold: both were from women aged 65 years or older, for whom one pyuria point is deducted.
\* No sample could be classified Definite UTI because the systemic domain scored zero for every sample, making 7 the maximum attainable total.

Of the 136 samples meeting the entry pyuria threshold conservatively, all 136 (61.3% of all samples) were classified Possible or Probable UTI; the 86 that failed this threshold were unscored. Of the 206 samples meeting the entry pyuria threshold inclusively, 204 (91.9% of all samples) were classified as Possible or Probable UTI. Every sample from someone with SCI/D carries one symptom point and zero systemic points, so a sample meeting the pyuria threshold enters scoring with a total of at least 2 points. All but two of our 222 samples entered scoring with at least 3 points, putting them at the lower bound of Possible UTI before any culture result. The culture domain determines whether a sample is called Possible or Probable. The two exceptions were women aged ≥65 years, in whom the pyuria deduction reduced the total to 2.

Of the 222 samples, the individuals providing 192 samples remained asymptomatic for 72 hours after the sample, and after 30 samples the individuals developed at least one symptom on the USQNB-IC within 72 hours. Table 1 shows that Probable UTI was assigned to 38.5% and 40.0% of these samples, respectively under the conservative interpretation of pyuria bins and 51.0% and 53.3% under the inclusive interpretation. Whether individuals went on to develop symptoms or not did not affect the classifications.

The difference between the two interpretations arises in two distinct ways. Seventy samples (32.6%) were reported as “00-05”, which straddles the entry threshold: under the conservative reading all 70 exit as No UTI, under the inclusive reading all 70 enter scoring. That bin accounts for the whole of the difference in routing; no other bin changes whether a sample is evaluated. A second bin, “40-60”, straddles the boundary between 2 and 3 pyuria points and so changes grading rather than routing, moving 5 samples from Possible to Probable UTI. Overall, 73 of the 222 samples are classified differently under the two readings: 68 because they are evaluated at all, and 5 because they are graded differently once evaluated. Among samples entering scoring, the pyuria domain contributed almost no variance to the total (variance 0.125 conservative, 0.155 inclusive). Table 2 summarizes how points accrue by domain.

**Table 2.**
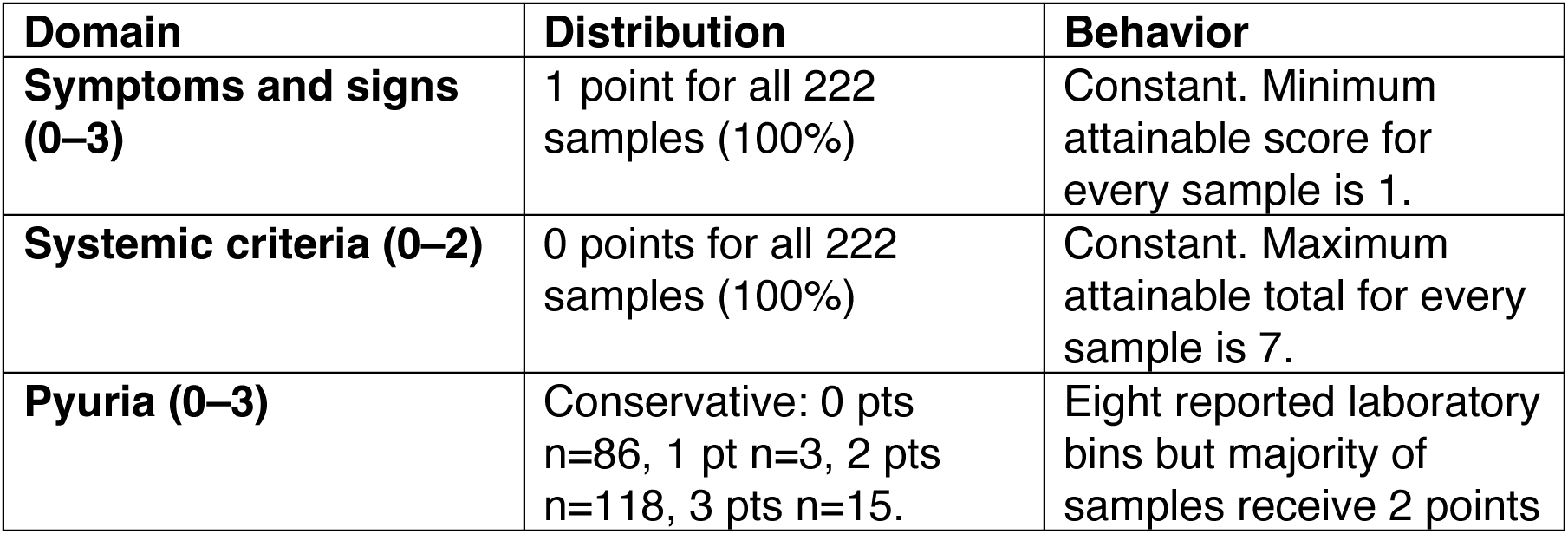
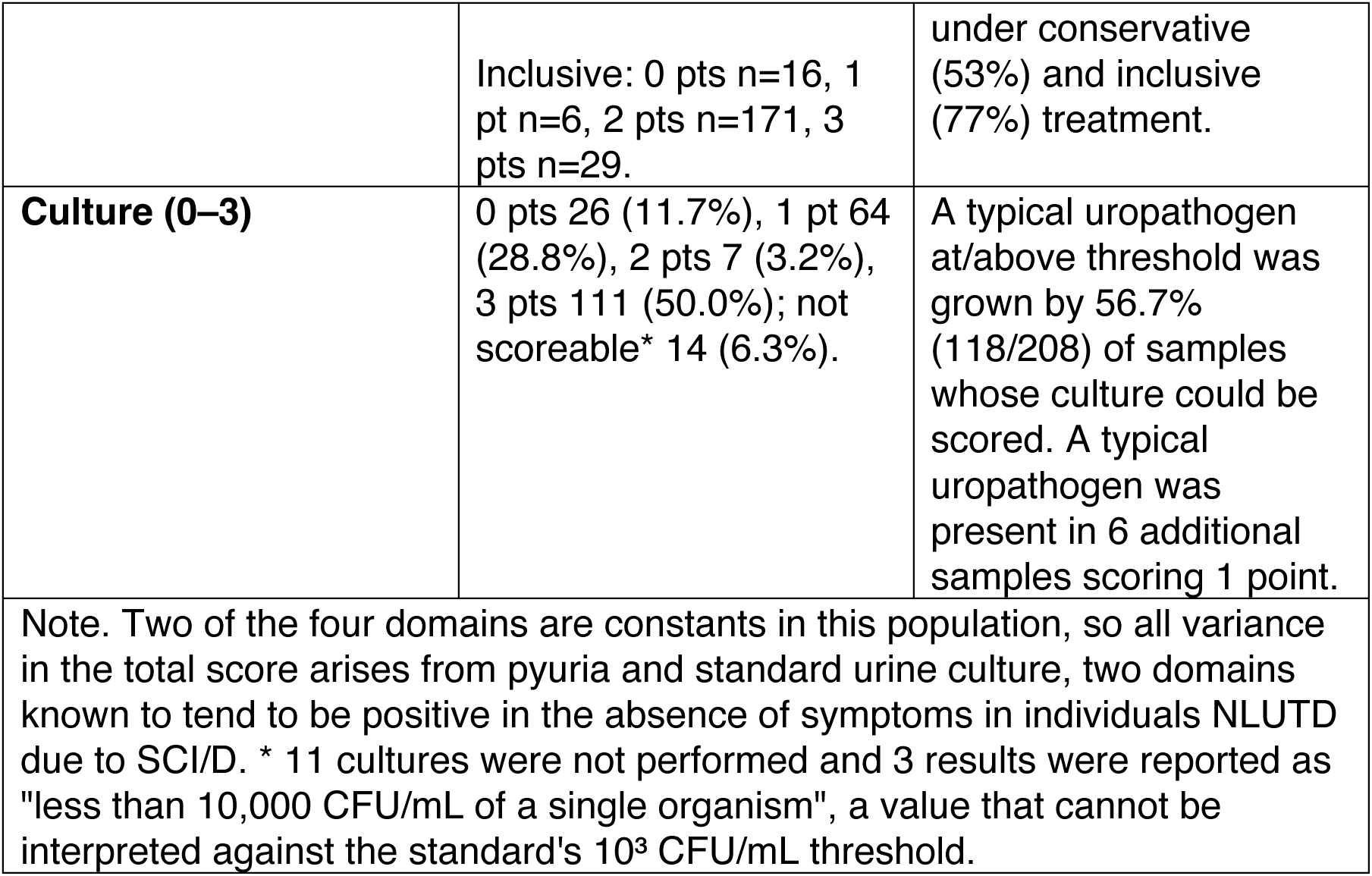
Behavior of the four scoring domains in 222 samples from 96 adults with SCI/D asymptomatic for 72 hours prior to sample.

Figure 2 shows the distribution of total scores. Totals of 0 are unattainable, since every sample receives one symptom point from the SCI diagnosis and any sample entering scoring receives at least one pyuria point; totals of 8 or more are unattainable because the systemic domain is zero throughout. The attainable range is therefore 2 to 7 for asymptomatic individuals with SCI/D.

**Figure 2.**
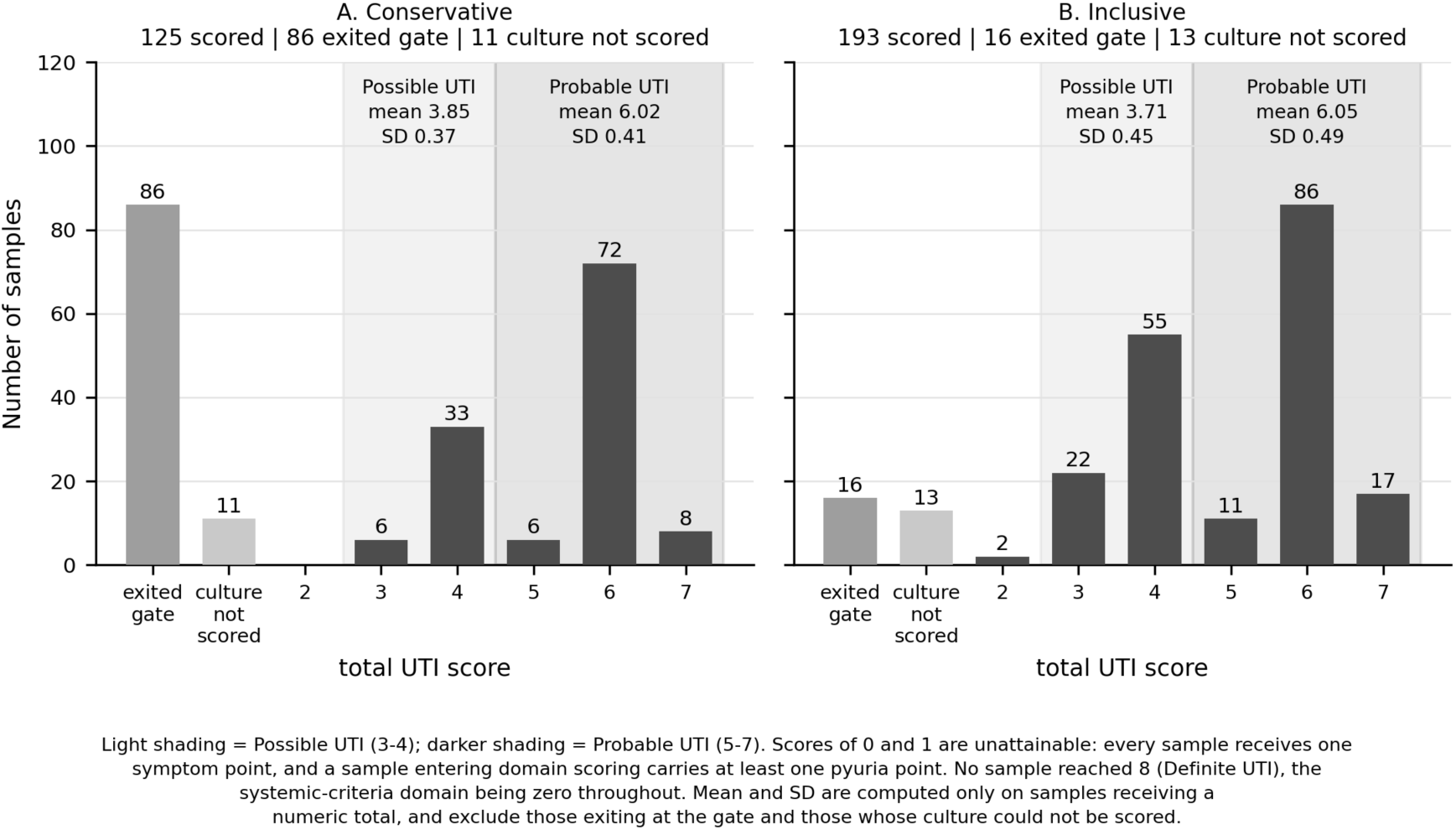
Distribution of total UTI scores under the conservative (A) and inclusive (B) interpretations. Light shading marks Possible UTI (3-4) and darker shading Probable UTI (5-7). Two categories of sample carry no numeric total and are shown as separate bars: those exiting at the pyuria entry threshold, and those whose culture could not be scored because it was never performed or was reported in a form the standard cannot grade. Totals of 0 and 1 are unattainable, every sample receiving one symptom point and any sample entering scoring at least one pyuria point; no sample reached 8 or more, the systemic domain scoring zero throughout.

A typical uropathogen at/above threshold was grown by 56.7% (118/208) of scorable culture results. However, what our laboratory reports is not consistent with what the standard requires: 208 of 222 cultures can be scored and 3 cannot (11 were not done). Only 178 were returned as a colony count. A further 30 were returned as a comment, rather than a quantitative result - 26 had a comment of “no growth”, and four had a comment about “mixed flora” and no quantification (see S2 Table).

Table 3 shows that, as expected in individuals with NLUTD due to SCI/D, both asymptomatic bacteriuria and pyuria were present together in the majority of these saples.

**Table 3.** Proportion of samples growing a typical uropathogen, by reported WBC bin, among the 211 samples on which a culture was performed (N = 211)*.

| Reported WBC bin** | n | % with typical uropathogen at any level |
| --- | --- | --- |
| None Seen | 16 | 56.2 |
| 00-05 | 69 | 42.0 |
| 06-10 | 37 | 51.4 |
| 10-20 | 30 | 73.3 |
| 20-40 | 31 | 83.9 |
| 40-60 | 14 | 78.6 |
| >60 | 13 | 53.8 |
| Packed | 1 | 100.0 |
| Note. * The denominator here is the 211 cultures performed, not the 208 that could be scored: whether an organism grew is determinable for the 3 samples reported as fewer than 10,000 CFU/mL of a single organism, even though the colony count cannot be placed relative to the standard's threshold and no culture points can be assigned.<br>** The clinical laboratory reports results as shown and does not allow alternative result categories. |  |  |

The proportion of samples with culture results including a typical uropathogen at any level increases as WBC per hpf increases to the “20-40” bin. The two domains are typically associated in asymptomatic individuals with SCI/D, so summing them counts a common characteristic of NLUTD twice, rather than combining independent evidence.

When the standard functions as a set of scoring rules rather than a routing algorithm, a sample could achieve zero pyuria points and proceed to culture (since many clinical protocols require both urinalysis and standard urine culture whenever one is ordered). Every sample carries one symptom point at entry, so a sample that scored zero on pyuria would still reach Possible UTI (3 points) on culture alone wherever culture contributed 2 or more points. That is the case for 35 of the 86 samples that exit conservatively and 9 of the 16 that exit inclusively, raising the proportion classified Possible or Probable to 171 of 222 (77.0%) and 213 of 222 (95.9%). Two ambiguities therefore act on the same result: how a reported bin that straddles a cut point should be read, and whether the pyuria criterion routes a sample or scores it. Across both, the proportion classified Possible or Probable UTI ranges from 61.3% to 95.9%, and neither ambiguity is resolvable from the text of the standard together with a routine laboratory report.

## Discussion

Applied to 222 urine samples from adults with NLUTD due to SCI/D using IC who confirmed they were asymptomatic for 72 hours prior to sample, the 2024 Bilsen consensus reference standard classified 38.7–51.4% as Probable UTI and a further 22.5–40.5% as Possible UTI, depending on the interpretation of categorical WBC boundaries. Under the inclusive interpretation, only 8.1% of these asymptomatic samples were classified No UTI.

The mechanism is structural. Every sample receives one symptom point and zero systemic points, so any sample meeting the pyuria entry threshold carries 2 to 4 points before culture contributes - 3 points for 118 of the 136 samples scored under the conservative reading, and 2 only where the age-65 deduction applies. Baseline pyuria is common in this population in the absence of symptoms,4,5 and the organisms earning the highest culture scores are the conventional uropathogens that are common in the urobiome associated with SCI/D.8,11 The rates of Probable and Possible UTI classification are a floor: the systemic domain which we could not assess in our sample, contributed nothing because no participant was febrile and no bloods were drawn, which is routine for care of individuals with NLUTD due to SCI/D. The presence of pyuria and bacteriuria in this population is well known, but individuals with SCI are explicitly included in this UTI standard for research in spite of that.

A reference standard intended for research and for diagnosis must be scoreable from what laboratories actually issue. Ours issues ordered categorical bins rather than counts, ranges rather than values, free text in place of a result when nothing grew or too much did, and an empty organism field for cultures the standard expects to score. This particular lab does not enumerate growth that does not meet its internal standard for “clinically important”, and it does not perform every culture ordered. None of this is error - a clinical laboratory reports what it judges clinically actionable.

Conversely, a research laboratory might be more likely to report what the protocol specifies, for every specimen. The standard is written as though such a laboratory will process the samples: it requires a leukocyte count, a colony count against 10³ CFU/mL, and the identity of every organism. The categorical statements a clinical laboratory supplies are mostly usable - 208 of our 222 cultures could be scored. If the standard can be scored only where a research laboratory processes the urine, that is an unstated scope limitation, and a consequential one: it excludes the routine-care settings in which UTI in NLUTD due to SCI/D is actually diagnosed and over-treated. Investigators should establish in advance what their laboratory will report, and state how absent counts were treated.

At issue for clinical and research labs alike is a function of measurement capability. The entry threshold for pyuria separates 5 from 6 leukocytes per high power field, and the 2-to-3-point boundary separates 49 from 50 leukocytes per high power field. How precisely either can be measured depends on how leukocytes are counted, which varies between laboratories. Moreover, the framework permits two units of measurement for pyuria but does not treat them consistently: at entry it equates ≥10 per µL with ≥5 per hpf, a ratio of 2:1, and at the 2-to-3-point pyuria boundary, ≥200 per µL is equated with ≥50 per hpf, a ratio of 4:1. No single conversion satisfies both. A specimen between 100 and 200 per µL will therefore score 2 or 3 points according to which unit the laboratory reports.

Table 4 sets out the provisions of the standard that could not be applied as written, together with what routine practice supplies in their place. The subsections that follow take them in the order the algorithm applies them.

**Table 4.** Provisions of the reference standard that cannot be applied as written in this setting.

| Provision | What routine practice supplies | Consequence for the score |
| --- | --- | --- |
| **Question 1: capability to perceive or express symptoms** | Patient-reported symptoms collected with an instrument validated in this population<sup>12</sup>; SCI is named by the standard as an example of inability. | If answered based solely on diagnosis (yes/no SCI), the same fact removes the symptom-based exit and awards a point. |
| **Pyuria entry threshold, 5 per hpf** | Ordered categorical bins, one of which ("00-05", 70 samples) straddles the threshold. | Determines classification. Also may require discrimination outside the resolution of the counting method. |
| **Pyuria grading boundary, 50 per hpf** | A reported bin ("40-60") straddles the boundary. | Point assignment within the domain is not fully determinable from the laboratory report. |
| **Two permitted pyuria units** | A laboratory reports one unit or the other, not both. | The standard equates $10/\mu\text{L}$ with 5/hpf and $200/\mu\text{L}$ with 50/hpf - ratios of 2:1 and 4:1. Specimens between 100 and 200 per $\mu\text{L}$ score 2 or 3 points depending on the unit reported. |
| **Culture threshold, $\geq 10^3$** | Reported growth, which in this cohort cleared the | The threshold does not discriminate in this population. |
| **CFU/mL for a single catheterised specimen** | threshold in most of our cases. |  |
| **"Other pathogen", undefined** | Named organisms of widely differing significance, including skin and genital flora. | Organisms the laboratory regards as contaminants score the same as *Enterococcus* does. |
| **Zero-point culture cell** | A single reported value covering no growth, culture not performed, and growth the laboratory declined to characterise (37 samples). | A result assessed and negative and a missing result contribute identically to the total. |
| **Exclusion of catheter-associated UTI, undefined** | Clinical and surveillance definitions disagree about whether people with SCI/D using intermittent catheterization can use this reference standard or are excluded. | Whether the standard applies to this population will depend on the definition of CAUTI. |

The classification turns almost entirely on the first routing question, and the framework answers that question for the clinician. Our participants are demonstrably able to report urinary symptoms: they did so at every collection, and after 30 samples they reported symptom onset within 72 hours. The framework nonetheless names spinal cord injury as an example of inability, so a clinician unfamiliar with SCI/D will reasonably answer from the diagnosis, which removes the symptom-based exit and awards a point. Routing and scoring are therefore not separable, and the consequence is stark. Answered as the framework directs, 61.3-91.9% of these samples are Possible or Probable UTI. Answered from symptom data validated for this population13, every one of the 222 exits as No UTI, with no domain scored. Yet those same samples, had they been scored rather than routed out, would have been Possible or Probable UTI in 77.0 - 95.9% of cases. The same specimen, with the same laboratory values, is No UTI or Probable UTI according to which of two defensible readings of a single question a clinician makes, and the standard offers no way to choose between them.

Consensus methods aggregate expert judgment reliably, but rigorous conduct guarantees only that the output represents the panel’s beliefs, not that those beliefs hold in populations where they have not been validated. The thresholds are, on the authors’ own account, imported rather than validated: the systemic domain cutoffs were chosen “by extrapolating data from studies investigating UTI-related bloodstream infection and sepsis”, and cutoffs were provided chiefly “to ensure uniformity”.

Bilsen et al. concluded that CAUTI requires a separate standard because symptoms and markers are especially difficult to interpret there. These results demonstrate that the same grounds apply for individuals with NLUTD due to SCI/D. Clinical definitions of catheter-associated UTI include intermittent catheterization14,15; surveillance definitions16 exclude it. The framework’s culture domain follows the clinical definition, specifying ≥10³ CFU/mL for a single catheterized specimen, while its symptoms domain adopts surveillance structure. Under a clinical reading this cohort is included and named; under a surveillance reading and based on the conclusion that CAUTI requires a separate standard, this standard was expected to fail as systematically as it did.

The implications extend beyond SCI/D. Urinary marker unreliability is increasingly recognized, because the underlying premise - that pyuria and uropathogen growth mean infection - rests on the now-falsified assumption that urine is sterile.8 This recognition has not prevented its opposite being codified, as this 2024 standard has done. Criteria calibrated in adults without NLUTD cannot be assumed to transfer to any condition that alters urinary biology at baseline. We have also identified more general challenges in applying the standard, including the difficulty in reconciling domain thresholds with our (and probably others’) laboratory’s result reporting, the inseparability of routing from scoring, and thresholds set below the resolution of the measurement.

Limitations. This is a secondary analysis of samples from individuals recruited from a single site. No blood markers were collected so we could not assess the systemic domain, and no individuals without NLUTD were included who might have supported an analysis focused on CAUTI among those using intermittent catheterization on a temporary basis. Laboratory reporting was categorical and reflects one laboratory’s conventions. Samples are clustered within participants, and the domain audit was descriptive rather than prespecified. However, our repeated measures, 72 hour windows for participant confirmed (prior) and USQNB-IC verified (post) asymptomatic status, and large sample size strengthen confidence in our conclusions.

## Conclusions

Pyuria in NLUTD due to SCI/D is produced by the bladder dysfunction and its management rather than by infection, and the organisms that score highest are those that populate the urobiome in this population. A scoring algorithm encodes a causal model whether or not that model is stated: summing domains treats each as an indicator of one condition and their contributions as exchangeable. That is a substantive causal claim, and here it fails.

We suggest that future consensus criteria state, for each component, what causal relation to the target condition is assumed, in which population it has been established, and what plausible alternative causes could produce that component. Stating the model makes it checkable. AGREE II17 appraises how a guideline was developed rather than the validity of what was developed, and GRADE18 rates certainty in evidence rather than the construct validity of consensus-derived criteria.

For UTI research in community-dwelling individuals with SCI/D and NLUTD, this analysis does not support the use of the 2024 research reference standard: its scoring pathway cannot return “No UTI” for asymptomatic samples in the presence of pyuria and bacteriuria in this population.

## Contributors

*RET designed the secondary analysis, applied the reference standard scoring algorithm to the samples, verified the scoring against the source laboratory records, prepared the tables and figures, and drafted the manuscript. SLG contributed to interpretation of the findings and to critical revision of the manuscript. Both authors had full access to the data reported here, contributed to the decision to submit, and read and approved the final version. Both authors meet the ICMJE criteria for authorship and accept responsibility for the integrity of the analysis*.

## Declaration of interests

*Both authors declare intellectual interests in the subject matter of this manuscript. RET and SLG developed the Urinary Symptom Questionnaire for Neurogenic Bladder (USQNB) instruments, validated separately for intermittent catheter, indwelling catheter and voiding bladder management, which we cite as evidence that symptom report is obtainable in this population; they also developed the cUTI likelihood profiling framework and led the revision of the International Spinal Cord Society UTI Basic Data Set, both of which the Discussion identifies as candidate population-appropriate standards. None of these is commercial: the instruments are freely available and neither author receives royalties, licensing income or any other payment arising from their use. We have not compared the reference standard evaluated here against any of these instruments; our claims concern only whether the 2024 standard can be applied as written in this population. Readers should weigh our conclusions in that light. SLG declares a financial relationship with, and intellectual property holdings in, Uvantis Therapeutics, Inc., unrelated to the submitted work; and employment by MedStar National Rehabilitation Hospital. RET declares employment by Georgetown University; paid consultation on a Data Safety Monitoring Board for the Kessler Foundation unrelated to this work; paid service on the Patient-Centered Outcomes Research Institute’s Methodology Committee unrelated to this work; royalties derived from two books on ethical statistical and data science practice unrelated to this work; a research fellowship (unpaid) at MedStar National Rehabilitation Hospital; an unpaid role on the Committee on Professional Ethics of the Association for Computing Machinery; an unpaid role on the Advisory Board on Ethics of the International Statistical Institute; an unpaid role on the UK National Statisticians Data Ethics Committee; and an unpaid role as a section editor for PLOS ONE. None of these organizations had a role in this manuscript. Both authors received salary support from Department of Defense grant W81XWH-19-1-0541, which funded collection of the data analysed here; the funder had no role in study design, data collection, analysis, interpretation, or the decision to submit. Beyond these, neither author has any financial or personal relationship with any person or organisation, within the three years preceding the work submitted, that could be perceived to influence it: no consultancies, stock ownership, honoraria, paid expert testimony, patents or patent applications, or travel grants*.

## Data availability

*De-identified individual participant data supporting this analysis are available from Zenodo at doi:10.5281/zenodo.21909574. The deposit provides one row per urine sample, with the symptom record, the reported white blood cell bin, the reported culture result and its classification, and the pyuria and culture domain points under both the conservative and inclusive readings, so that every classification reported here can be reproduced. Age, diagnosis, time since injury and level of injury are withheld: released at the individual level in a cohort of this size they would render participants identifiable. Age enters the reference standard only through the deduction applied to women aged 65 years or older, which affects 9 samples from 4 participants; that deduction is reproducible from a released Boolean indicator rather than from age itself*.

## Supporting information

S1 Table. What the laboratory reported for pyuria, and where each reported bin sits relative to the reference standard’s cut points. Gives all of our laboratory’s eight white blood cell bins with their frequencies, the points each is given under both readings, and which bins straddle a cut point or are not counts at all.

**S1 Table.** What the laboratory reported for pyuria, and where each reported bin sits relative to the reference standard’s cut points (N = 222).

| Reported white blood cell result | n | Where it sits relative to the standard's cut points | Points, conservative | Points, inclusive |
| --- | --- | --- | --- | --- |
| "None Seen" | 16 | NOT A COUNT. Taken as below the entry threshold of 5 per hpf. | 0 - exits | 0 - exits |
| "00-05" | 70 | STRADDLES the entry threshold: the bin contains values below 5 and the value 5 itself. These 70 samples are the whole of the difference between the two readings. | 0 - exits | 1 or 2 - enters |
| "06-10" | 42 | Determinable: entirely above the entry threshold and entirely below the 50 per hpf boundary. | 2 | 2 |
| "10-20" | 31 | Determinable, as above. | 1 or 2 | 1 or 2 |
| "20-40" | 32 | Determinable, as above. | 2 | 2 |
| "40-60" | 16 | STRADDLES the boundary between 2 and 3 points at 50 per hpf. | 1 or 2 | 2 or 3 |
| ">60" | 13 | Determinable: entirely above the 50 per hpf boundary. | 3 | 3 |
| "Packed" | 2 | NOT A COUNT. Taken as above the 50 per hpf boundary (>60). | 3 | 3 |

**Notes.** The standard requires a leukocyte count in either of two units. This laboratory reports ordered categorical bins. Two of the eight straddle a cut point the standard uses, and two are not counts at all. No sample could be assigned its pyuria points from the report alone without a decision - taken by the investigator, not by the standard - about how to read a straddling bin. Where two point values are shown, the lower arises from
the subtraction of one point for women aged 65 years or older, which applies to 9 samples from 4 participants.

S2 Table. What the laboratory reported for urine culture, and the scoring problem each form of report creates. Gives each form in which a result was returned, its frequency, the points assigned, and whether the result is scoreable, scoreable but recoverable only from a free-text field, or not determinable at all.

**S2 Table.**
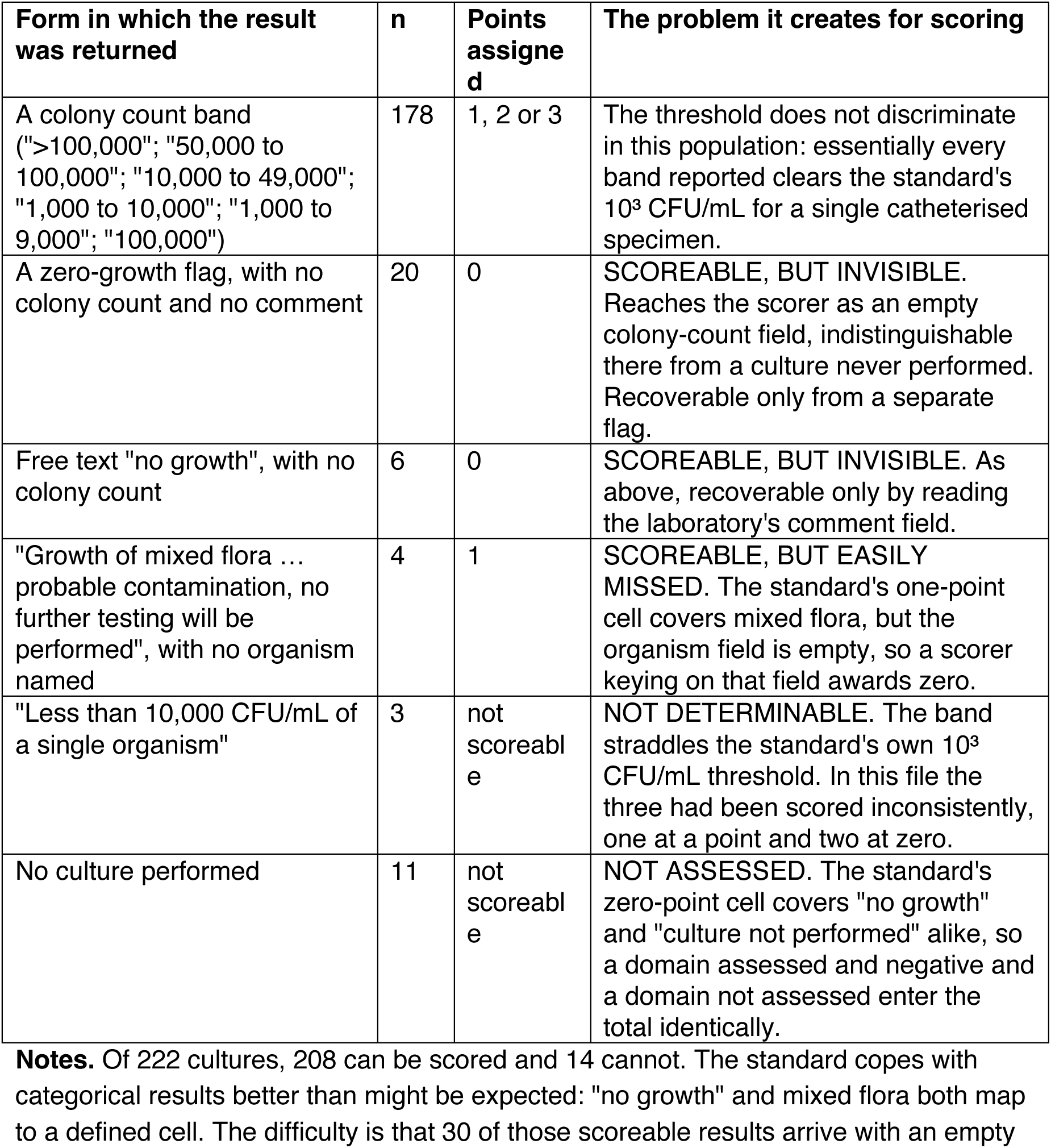
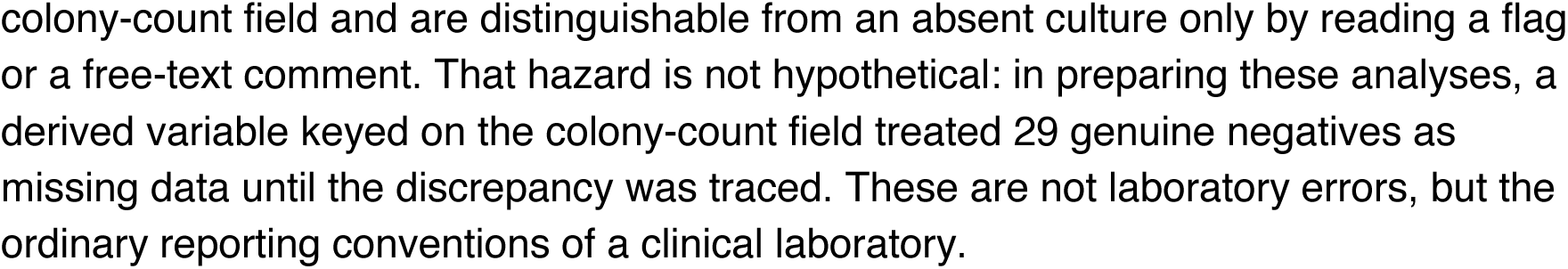
What the laboratory reported for urine culture, and the scoring problem each form of report creates (N = 222).

| Form in which the result was returned | n | Points assigned | The problem it creates for scoring |
| --- | --- | --- | --- |
| A colony count band (">100,000"; "50,000 to 100,000"; "10,000 to 49,000"; "1,000 to 10,000"; "1,000 to 9,000"; "100,000") | 178 | 1, 2 or 3 | The threshold does not discriminate in this population: essentially every band reported clears the standard's 10<sup>3</sup> CFU/mL for a single catheterised specimen. |
| A zero-growth flag, with no colony count and no comment | 20 | 0 | SCOREABLE, BUT INVISIBLE. Reaches the scorer as an empty colony-count field, indistinguishable there from a culture never performed. Recoverable only from a separate flag. |
| Free text "no growth", with no colony count | 6 | 0 | SCOREABLE, BUT INVISIBLE. As above, recoverable only by reading the laboratory's comment field. |
| "Growth of mixed flora ... probable contamination, no further testing will be performed", with no organism named | 4 | 1 | SCOREABLE, BUT EASILY MISSED. The standard's one-point cell covers mixed flora, but the organism field is empty, so a scorer keying on that field awards zero. |
| "Less than 10,000 CFU/mL of a single organism" | 3 | not scoreable | NOT DETERMINABLE. The band straddles the standard's own 10<sup>3</sup> CFU/mL threshold. In this file the three had been scored inconsistently, one at a point and two at zero. |
| No culture performed | 11 | not scoreable | NOT ASSESSED. The standard's zero-point cell covers "no growth" and "culture not performed" alike, so a domain assessed and negative and a domain not assessed enter the total identically. |

**Notes.** Of 222 cultures, 208 can be scored and 14 cannot. The standard copes with categorical results better than might be expected: "no growth" and mixed flora both map to a defined cell. The difficulty is that 30 of those scoreable results arrive with an empty
colony-count field and are distinguishable from an absent culture only by reading a flag or a free-text comment. That hazard is not hypothetical: in preparing these analyses, a derived variable keyed on the colony-count field treated 29 genuine negatives as missing data until the discrepancy was traced. These are not laboratory errors, but the ordinary reporting conventions of a clinical laboratory.

## Notes

This work was supported by Department of Defense grant # W81XWH-19-1-0541. The funders had no role in study design, data collection and analysis, decision to publish, or preparation of the manuscript.

### Competing Interest Statement

RET developed the cUTI likelihood profiling approach and led the ISCoS UTI Basic Data Set revision, both of which this analysis identifies as the population-appropriate alternative. RET and SLG developed the USQNB instruments referenced herein. SLG is developing an application operationalizing the cUTI approach. Additional interests are described at the end of the manuscript.

### Author Declarations

IRB of MedStar National Rehabilitation Hospital gave ethical approval for this work (#1124)

### Summary of Updates

We have expanded the analyses of the 2024 UTI research standard and the manuscript itself which is now roughly 3 times longer and includes figures and tables. We also corrected errors in the original data file and verified the scoring. This manuscript is in review and once finalized, the data will be published in Zenodo (DOI in progress).

